# Prognostic value of plasma IgG N-glycome traits in patients with pulmonary arterial hypertension: an observational cohort study

**DOI:** 10.1101/2023.12.04.23299348

**Authors:** Ze-Jian Zhang, Chao Liu, Jie-Ling Ma, Jing-Si Ma, Jia Wang, Ruo-Nan Li, Dan Lu, Yu-Ping Zhou, Tian-Yu Lian, Si-Jin Zhang, Jing-Hui Li, Lan Wang, Kai Sun, Chun-Yan Cheng, Wen-Hui Wu, Zhi-Cheng Jing

## Abstract

**Background:** BNP (brain natriuretic peptide) or NT-proBNP (N-terminal proBNP) is the only blood biomarker in the established risk calculators for pulmonary arterial hypertension (PAH). New plasma biomarkers of systemic origin may improve the risk assessment in PAH.

**Methods:** We utilized a robust mass spectrometry-based method to identify prognostic plasma IgG N-glycome traits in PAH patients from two national referral centers in China (Beijing [discovery] cohort, n = 273; Shanghai cohort [validation], n = 349). Then the candidate IgG N-glycan traits were evaluated in combined cohorts adjusted for potential confounders and subgroup analyses. At last, the candidate IgG N-glycan traits were further evaluated against the established prognostic risk equation (three-strata model from the SPAHR and COMPERA analyses) for PAH. The primary endpoint was all-cause mortality.

**Results:** IgG Fucosylation was found to predict survival independent of age and sex in the discovery cohort (HR: 0.377, 95% CI: 0.168-0.845, *P* = 0.018) with confirmation in the validation cohort (HR: 0.264, 95% CI: 0.445-0.751, *P* = 0.005). IgG Fucosylation remained a robust predictor of mortality in PAH patients in the combined cohorts adjusting for potential confounders and in subgroup analyses based on sex and treatment strategies. Moreover, the addition of the IgG Fucosylation to the established three-strata model significantly improved the predictive accuracy from AUC of 0.67 to AUC of 0.70 (*P* = 0.043) and IgG Fucosylation was useful in further stratifying the intermediate risk PAH patients classified by the three-strata model into intermediate-low and intermediate-high risk subgroups.

**Conclusions:** The plasma IgG Fucosylation informs prognosis independent of the existing clinical assessments in PAH and may be useful in the clinical management of patients with PAH.

**Clinical Perspective What is new?:**

- The plasma IgG Fucosylation, which is in different pathophysiological pathways from other established risk factors of pulmonary arterial hypertension (PAH), can inform prognosis independent of the existing clinical assessments in patients with PAH.
- The addition of the IgG Fucosylation to the established three-strata model for PAH prognosis significantly improved the predictive accuracy.
- IgG Fucosylation was useful in further stratifying the intermediate risk PAH patients classified by the three-strata model into intermediate-high and intermediate-low risk subgroups.

**What are the clinical implications?:**

- Plasma IgG N-glycan biomarkers may provide a refined risk stratification in PAH used in combination with the established risk factors.
- IgG Fucosylation involved in inflammatory pathways may not only be a promising prognostic biomarker but also a potential novel therapeutic target, distinct from existing targets, for patients with PAH.

## Introduction

Pulmonary arterial hypertension (PAH) represents a rare and malignant cardiopulmonary condition characterized by dysregulation of endothelium-derived vasoactive factors, inflammatory processes, and obstructive remodeling of small pulmonary arteries. These pathological changes lead to a progressive increase in pulmonary vascular resistance (PVR) and mean pulmonary artery pressure, culminating in right heart failure and, ultimately, death for the majority of affected patients.^1,2^

The prognosis of PAH remains poor with an estimated 5-year survival rate of 60% for patients with advanced PAH.^3–5^ Nevertheless, treatment response varies between patients due to the heterogeneity of the disease,^6^ and life expectancy is quite variable. Risk stratification and regular assessment of disease severity are vital to guide the clinical management of patients with PAH.^1,2^ The existing guidelines recommend multiparametric risk calculators based on clinical assessment, right ventricular function, exercise, and hemodynamic variables to inform prognosis and guide treatment decision-making in PAH.^1,2^ However, the multidimensional approach suffers from some disadvantages and limitations, such as not all these measurements available at each patient visit, some factors obtained invasively, unsatisfied accuracy, and restricted mechanisms reflected by the parameters. Moreover, in the established multifactorial prognostic model, the circulating biomarker includes only BNP (brain natriuretic peptide) or NT-proBNP (N-terminal proBNP). Abnormal immune regulation and inflammation have been recognized as critical contributors to the pathogenesis of PAH.^7–9^ New plasma biomarkers of systemic origin reflecting different components of the pathophysiology of PAH such as immune dysregulation and inflammation may improve the risk assessment of PAH and provide a more sensitive measure of disease severity and therapeutic response.

Glycosylation is a prevalent and important post-translational modification that modulates a variety of biological functions.^10,11^ Abnormal glycosylation of proteins is thought to play a role in almost every major disease.^12,13^ IgG, the predominant glycoprotein and antibody in human biofluids, is a key effector of the humoral immune system and has multiple roles in maintaining the balance of inflammation at the systemic level.^14^ The conserved N-linked glycans bound to the unique glycosylation site Asn297 in the Fc domain of IgG are of the complex biantennary type.^15,16^ The N-glycan compositions on IgG, particularly the core fucose moiety and the terminal galactose or sialic acids, exhibit variability and hold pivotal roles in regulating IgG structural stability and functional activity, which influences the outcome of the immune response (e.g., antibody’s effector functions).^17^ For example, it has been widely acknowledged that decreased levels of IgG core fucose contribute to increased antibody-dependent cellular cytotoxicity (ADCC) and inflammation.^18–21^ IgG loss of terminal galactose could trigger a proinflammatory response.^22,23^ It is noteworthy that our team has previously demonstrated dysregulation of the plasma IgG N-glycome in patients with chronic thromboembolic pulmonary hypertension^24^ and the identified abnormal IgG N-glycan traits may play a crucial role in the pathogenesis and progression of this condition.^24^ Nevertheless, the glycosylation characteristics of IgG and their prognostic value in patients with PAH have not yet been explored.

The main objective of this study was to identify and validate plasma IgG N-glycan traits that predict survival in patients with PAH to improve risk stratification.

## Methods

### Study population

From October 2006 to December 2019, all consecutive patients with a definite diagnosis of idiopathic PAH or heritable PAH at two national pulmonary hypertension referral centres (Shanghai Pulmonary Hospital with patients mainly from South China and FuWai Hospital in Beijing with patients mainly from North China) were included in this study. Idiopathic PAH was diagnosed based on baseline right heart catheterization confirming newly diagnosed precapillary pulmonary hypertension, defined by a resting mean pulmonary artery pressure≥25 mmHg, pulmonary artery wedge pressure≤15 mmHg, and PVR>3 Wood units, in the absence of other known causes of PAH.^1,2^ Patients with PAH were diagnosed with heritable PAH if their first or second-degree relatives had a confirmed PAH diagnosis or died from a PAH-like syndrome.^1,2^ The ethics review boards of Shanghai Pulmonary Hospital and FuWai Hospital approved the protocols. Written informed consent was obtained from all patients. This study complied with the principles of the Declaration of Helsinki.

### Baseline blood samples, data collection, and follow-up

Baseline demographic and clinical characteristics, including World Health Organization functional class (WHO FC), 6-minute walking distance (6MWD), and invasive haemodynamics including right arterial pressure (RAP), mean pulmonary artery pressure, pulmonary artery wedge pressure, cardiac index, PVR, and mixed venous oxygen saturation (SvO_2_) were collected. Baseline venous plasma for IgG N-glycome profiling and haemodynamic data were obtained during the first diagnostic right heart catheterization. Follow-ups were carried out through clinic visits or phone or internet interviews, and the patient’s survival time was calculated by months from the initial PAH diagnosis date to January 2023 or the latest contact date. The primary clinical endpoint was all-cause mortality.

### Mass spectrometric IgG N-glycome profiling

The profiling of IgG N-glycome was carried out using matrix-assisted laser desorption/ionization time-of-flight mass spectrometry,^25–27^ with raw mass spectrometry data processing performed according to established methods outlined in previous reports.^25–27^ Detailed procedures are provided in the Supplementary Material for reference.

To combine the effects of individual detected N-glycan traits sharing similar structures and better interpret the biological effects of IgG N-glycosylation,^28,29^ derived (calculated) N-glycan traits representing the most important features of IgG N-glycosylation including Galactosylation, Bisecting N-acetylglucosamine, and Fucosylation were calculated from the directly detected N-glycans. The formulas used for the calculation: Galactosylation = H3N4F1/(H4N4F1 + 2*H5N4F1); Bisecting N-acetylglucosamine = H3N5F1 + H4N5F1 + H5N5F1; Fucosylation = H3N4F1 + H4N4F1 + H3N5F1 + H5N4F1 + H4N5F1 + H5N5F1 (H = hexose; N = N-acetylhexosamine; F = deoxyhexose [fucose]).^24,27^ GlycoWorkbench software (version 1.1.3480) was used to annotate the N-glycan structures.

### Statistical analysis

Normality was determined using the Kolmogorov-Smirnov test. Continuous variables were presented as mean ± SD for normally distributed data, and as median (interquartile range, 25% to 75%) for non-normally distributed variables. Categorical variables were presented as n (%). Statistical analysis was performed with Statistic Package for Social Science (SPSS Inc., Chicago, USA) version 22.0 and GraphPad Prism version 8.0.0 for Windows (GraphPad Software, San Diego, California USA, www.graphpad.com). All tests were two-tailed with a value of P<0.05 considered statistically significant.

Prognostic IgG N-glycan traits were identified by Cox regression analysis corrected for age and sex in the discovery cohort and confirmed in the validation cohort. To assess whether the prognostic IgG N-glycan trait screened out was an independent prognostic factor, Cox regression analysis adjusted for potential confounders including age, sex, type of PAH, treatment strategies, 6MWD, WHO FC, NT-proBNP, RAP, SvO_2_, and cardiac index were conducted. Further adjusted Cox regression analyses were conducted in subgroups stratified by sex and therapeutic strategies. To assess the added value of IgG N-glycan traits to the established risk model, we compared the prognostic accuracy of the three-strata model (the established prognostic risk equation derived from the SPAHR and the COMPERA registry)^30,31^ with that of the IgG N-glycan + three-strata model using receiver-operating characteristic (ROC) analysis. Additionally, IgG N-glycans were employed to further stratify PAH patients classified as intermediate risk by the three-strata model, utilizing Kaplan-Meier survival analysis. The optimal cut-off values for the prognostic IgG N-glycan traits in Kaplan-Meier and Cox regression analyses were determined based on the values that maximized the sum of sensitivity and specificity in ROC analysis.^32^

## Results

### Study population and baseline characteristics

The study design is illustrated in Figure 1. A total of 273 patients with PAH enrolled from Fuwai Hospital between 2013 and 2019 (discovery cohort) and 349 patients with PAH enrolled from Shanghai Pulmonary Hospital between 2006 and 2013 (validation cohort) were included in this study.

**Figure 1.**
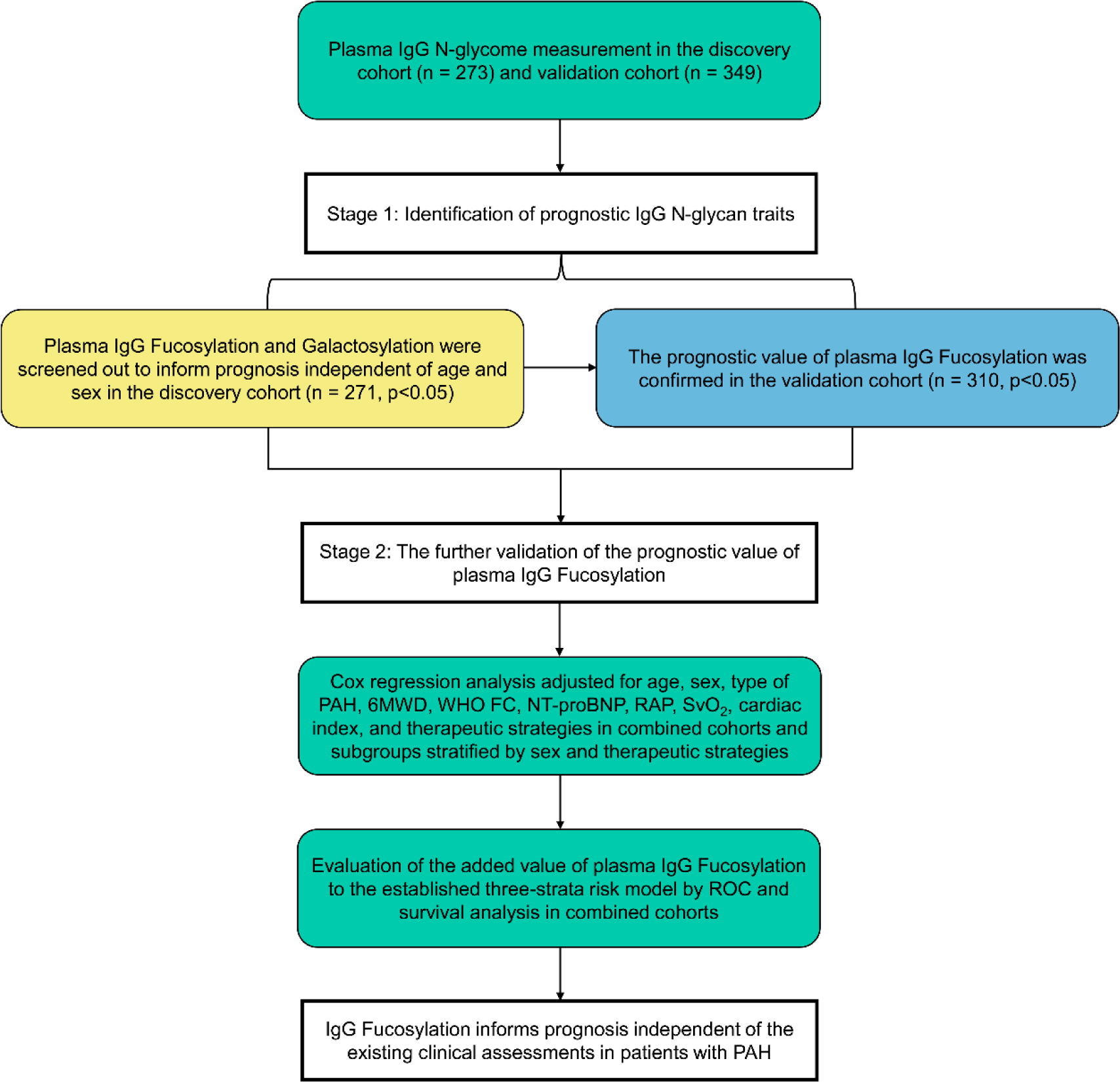
Study design. 6MWD = 6-minute walking distance; NT-proBNP = N- terminal pro-B-type natriuretic peptide; PAH = pulmonary arterial hypertension. PVR = pulmonary vascular resistance; RAP = right atrial pressure; ROC = receiver- operating characteristic; SvO_2_ = mixed venous oxygen saturation; WHO FC = World Health Organization functional class.

Among the patients, 58 (21.2%) in the discovery cohort and 109 (31.2%) in the validation cohort were male. The mean ages were 32.8 ± 10.4 and 35.9 ± 16.8 years in the discovery and validation cohorts, respectively. In the discovery cohort, 11 (4.0%) patients received untargeted therapy, 80 (29.3%) patients received monotherapy, 148 (54.2%) patients received dual therapy, and 34 (12.5%) patients received triple therapy. In the validation cohort, 66 (18.9%) patients received untargeted therapy, 227 (65.0%) patients received monotherapy, 53 (15.2%) patients received dual therapy, and 3 (0.9%) patients received triple therapy (Table 1).

**Table 1.** Baseline characteristics.

|  | **Discovery Cohort**<br>**(n = 273)** | **Validation Cohort**<br>**(n = 349)** |
| --- | --- | --- |
| Recruitment period | 2013-2020 | 2006-2013 |
| Age, y | 32.8 ± 10.4 | 35.9 ± 16.8 |
| Male, n (%) | 58 (21.2) | 109 (31.2) |
| Ethnic origin | Asian | Asian |
| IPAH/HPAH, n (%) | 259/14 (94.9/5.1) | 338/11 (96.8/3.2) |
| WHO FC, n (%) |  |  |
| I | 7 (2.8) | 6 (1.8) |
| II | 103 (41.2) | 107 (32.7) |
| III | 124 (49.6) | 199 (60.9) |
| IV | 16 (6.4) | 15 (4.6) |
| 6MWD, m | 416.8 ± 103.0 | 377.4 ± 116.7 |
| NT-proBNP, pg/mL | 941.0 (230.5–2,212.0) | 881.0 (289.0–1,944.0) |
| RAP, mm Hg | 8.4 ± 6.0 | 8.0 ± 5.5 |
| mPAP, mm Hg | 58.8 ± 15.6 | 61.4 ± 14.6 |
| PAWP, mm Hg | 9.5 ± 2.9 | 8.3 ± 3.3 |
| PVR, Wood units | 13.2 ± 6.1 | 15.4 ± 7.8 |
| Cardiac index, L/min/kg/m<sup>2</sup> | 2.5 ± 0.7 | 2.5 ± 0.9 |
| Cardiac output, L/min | 4.2 ± 1.3 | 4.0 ± 1.5 |
| SvO<sub>2</sub>, % | 65.5 ± 9.1 | 62.4 ± 10.8 |
| Three-strata risk (I/II/III), n (%) | 88/138/30 (33.1/55.6/11.3) | 96/206/37 (28.5/60.5/11.0) |
| Targeted therapy, n (%) |  |  |
| Treatment naive | 11 (4.0) | 66 (18.9) |
| Monotherapy | 80 (29.3) | 227 (65.0) |
| Dual therapy | 148 (54.2) | 53 (15.2) |
| Triple therapy | 34 (12.5) | 3 (0.9) |

6MWD = 6-minute walking distance; HPAH = heritable pulmonary arterial

### Follow-up

During a median follow-up of 39 (22-63) months, 41 patients (6.6%, 2 in the discovery cohort and 39 in the validation cohort) were lost to follow-up. In total, 139 patients (n = 52 in the discovery cohort and n = 87 in the validation cohort) died at the end of the follow-up periods.

### Plasma IgG N-glycome profiles in patients with PAH

Eight IgG N-glycans were directly detected in our cohorts by matrix-assisted laser desorption/ionization time of flight mass spectrometry. The representative annotated mass spectrometry spectra of the IgG N-glycome profiles from the survivor and non-survivor in our PAH cohorts are provided in Supplemental Figure S1, which demonstrates variations in peak patterns between the two groups. As derived N-glycan traits represent glycosylation changes shared by a group of structurally related N-glycans and enable better interpretation of the biological effects of glycosylation than directly detected N-glycan traits, three derived N-glycan traits including Galactosylation, Bisecting N-acetylglucosamine, and Fucosylation were calculated from the eight directly detected IgG N-glycans. We mainly evaluated IgG derived N-glycan traits in the present study.

### Identification of prognostic IgG N-glycan traits in patients with PAH

Discovery and validation cohorts were used to evaluate the associations of IgG N-glycan traits with the survival of patients. In the discovery cohort, Galactosylation (HR: 2.257, 95% CI: 1.274–4.001, *P* = 0.005; Figure 2A) and Fucosylation (HR: 0.377, 95% CI: 0.168–0.845, *P* = 0.018; Figure 2A) were found to predict survival independent of age and sex. Bisecting N-acetylglucosamine had no prognostic value (HR: 0.617, 95% CI: 0.219-1.737, *P* = 0.361; Figure 2A). We continued to validate our results in the validation cohort. However, only Fucosylation was confirmed to be associated with all-cause death independent of age and sex (HR: 0.264, 95% CI: 0.445–0.751, *P* = 0.005; Figure 2B) and the prognostic effect of Galactosylation was not replicated (HR: 1.092, 95% CI: 0.694–1.717, *P* = 0.705; Figure 2B). The Bisecting N-acetylglucosamine remained of no prognostic value in the validation cohort (HR: 1.789, 95% CI: 0.554–5.776, *P* = 0.330; Figure 2B). Finally, IgG Fucosylation was identified as a promising candidate prognostic glycan biomarker, which was further validated below.

**Figure 2.**
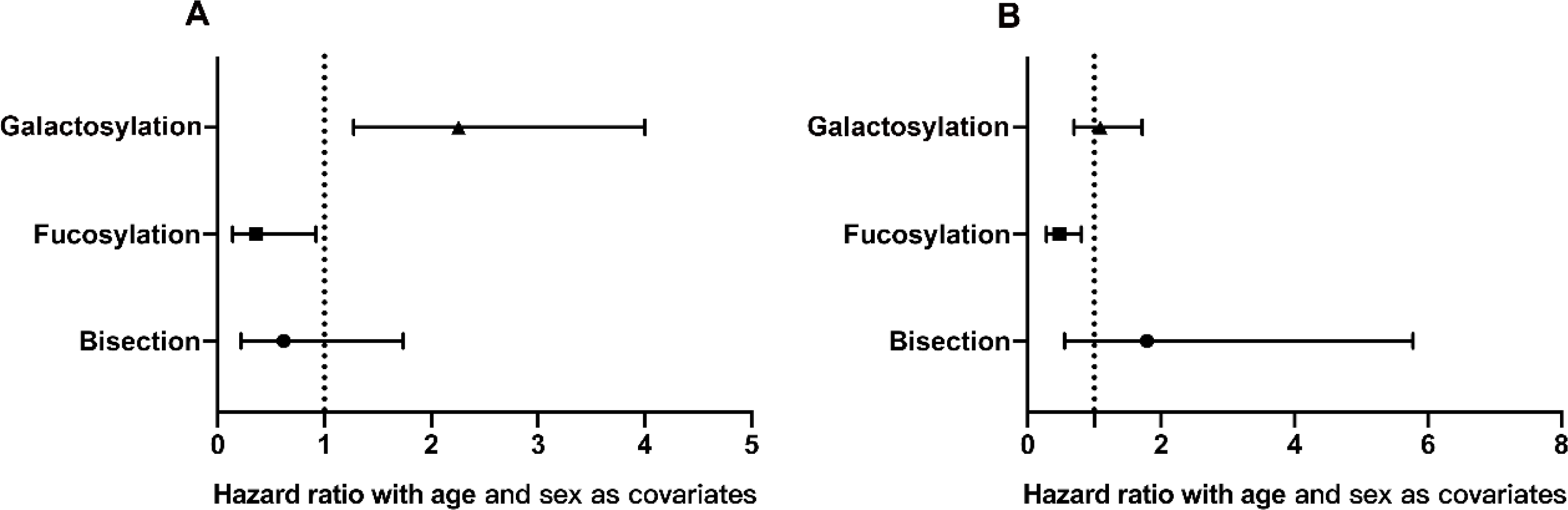
Hazard ratios and 95% CI from Cox regression analysis evaluating the prognostic IgG N-glycan traits in the discovery and validation cohorts.

### The validation of the prognostic value of IgG Fucosylation

We further evaluated the prognostic significance of IgG Fucosylation in the combined cohorts. In total, death occurred in 13.1% (25 out of 191) and 29.2% (114 out of 390) of patients in the high and low IgG Fucosylation groups, respectively, resulting in respective 5-year survival rates of 88.5% and 74.6% (*P* < 0.001 for the log-rank test; Figure 3).

**Figure 3.**
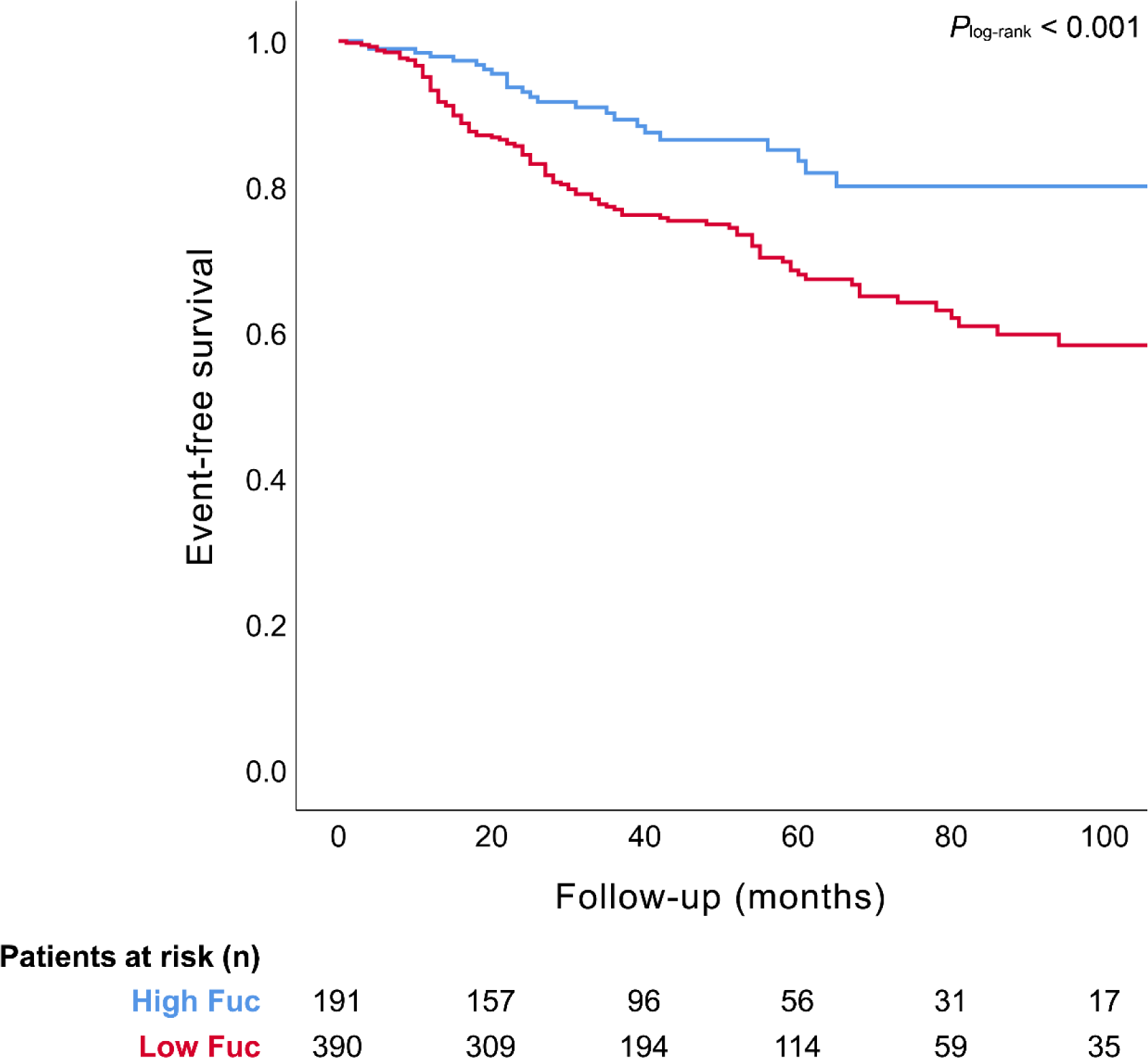
Kaplan-Meier survival curve for all-cause death stratified by IgG Fucosylation in patients with pulmonary artery hypertension. “High Fuc” or “Low Fuc” represents IgG Fucosylation higher or lower than 0.9741.

To evaluate whether IgG Fucosylation independently impacts the outcome of patients with PAH, Cox regression models were further adjusted for potential confounders including 6MWD, WHO FC, NT-proBNP, RAP, SvO_2_, cardiac index, type of PAH, and treatment strategies in addition to age and sex. Our results showed that the low IgG Fucosylation was still significantly associated with a high risk of all-cause death in patients with PAH (HR: 0.490; 95% CI: 0.315–0.763; *P* = 0.002; Table 2; Figure 4).

**Figure 4.**
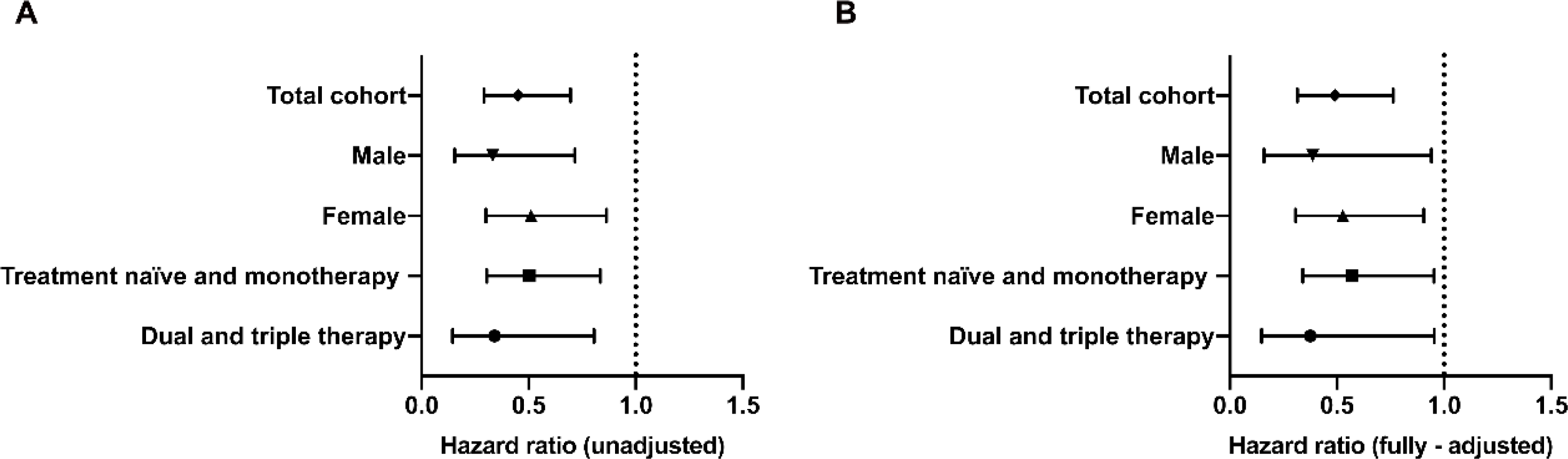
Hazard ratios and 95% CI from Cox regression analysis evaluating the IgG Fucosylation in combined cohorts and subgroup analyses (A) without adjustment, and (B) with full adjustment. The factors in the fully-adjusted models included age, sex, WHO-FC, 6MWD, NT- proBNP, RAP, SvO_2_, PVR, cardiac index, type of PAH, and therapeutic strategies. 6MWD = 6-minute walking distance; NT-proBNP = N- terminal pro-B-type natriuretic peptide; PAH = pulmonary arterial hypertension. PVR = pulmonary vascular resistance; RAP = right atrial pressure; SvO_2_ = mixed venous oxygen saturation; WHO FC = World Health Organization functional class.

**Table 2.** Results of Cox proportional hazards models examining the relationship between plasma IgG Fucosylation and risk of all-cause death in overall analysis and subgroup analyses according to sex and therapeutic strategies.

|  | Unadjusted |  | Fully-adjusted |  |  |
| --- | --- | --- | --- | --- | --- |
|  | HR (95%CI) | *P* value | HR (95%CI) | *P* value | *P* for interaction |
| Total (n = 581) | 0.451 (0.292-0.696) | <0.001 | 0.490 (0.315-0.763) | 0.002 |  |
| Sex |  |  |  |  | 0.748 |
| Male (n = 155) | 0.332 (0.154-0.715) | 0.005 | 0.387 (0.159-0.941) | 0.035 |  |
| Female (n = 426) | 0.510 (0.301-0.863) | 0.012 | 0.527 (0.307-0.906) | 0.014 |  |
| Therapy |  |  |  |  | 0.410 |
| Treatment naïve and monotherapy (n = 373) | 0.503 (0.304-0.834) | 0.008 | 0.570 (0.340-0.953) | 0.032 |  |
| Dual and triple therapy (n = 208) | 0.340 (0.144-0.805) | 0.014 | 0.376 (0.148-0.954) | 0.043 |  |

Furthermore, in order to comprehensively demonstrate the prognostic value of IgG Fucosylation, we further conducted subgroup analyses based on sex and therapeutic strategies. Our results showed that even after stratifying by sex, the association between low IgG Fucosylation and increased all-cause mortality remained robust and there was no significant interaction between sex and IgG Fucosylation (*P*= 0.784; Table 2); Interestingly, low Fucosylation posed a greater risk for males (fully adjusted models; male: HR: 0.387, 95% CI: 0.159–0.941, *P* = 0.035 and female: HR: 0.527, 95% CI: 0.307–0.906, *P* = 0.014; Table 2; Figure 4). Furthermore, we did not observe interactions between IgG Fucosylation and therapeutic strategies (*P*= 0.410; Table 2). We found that IgG Fucosylation continued to independently predict survival within subgroups stratified based on therapeutic strategies (fully adjusted models; dual and triple therapy: HR: 0.376, 95% CI: 0.148–0.954, *P* = 0.043; treatment naïve and monotherapy: HR: 0.595, 95% CI: 0.354–1, *P* = 0.033; Table 2; Figure 4).

At last, we investigated whether the IgG Fucosylation offered improvements in risk prediction in addition to the established prognostic risk equation (three-strata model from the SPAHR and COMPERA analyses)^30,31^ in PAH. We found the addition of IgG Fucosylation to the three-strata model improved the prediction of outcomes from area under the curve (AUC) 0.67 (95%CI: 0.62–0.72) to 0.70 (95%CI: 0.65–0.75) by ROC analysis (*P* = 0.043 for the difference in AUC) (Figure 5). In addition, about 60% of patients in our cohort met intermediate risk criteria in the tree-strata model (Figure 6A) which was consistent with previous reports, and IgG Fucosylation was useful in further dividing patients in the intermediate risk group into intermediate-high and intermediate-low risk subgroups (*P* < 0.001, Figure 6B).

**Figure 5.**
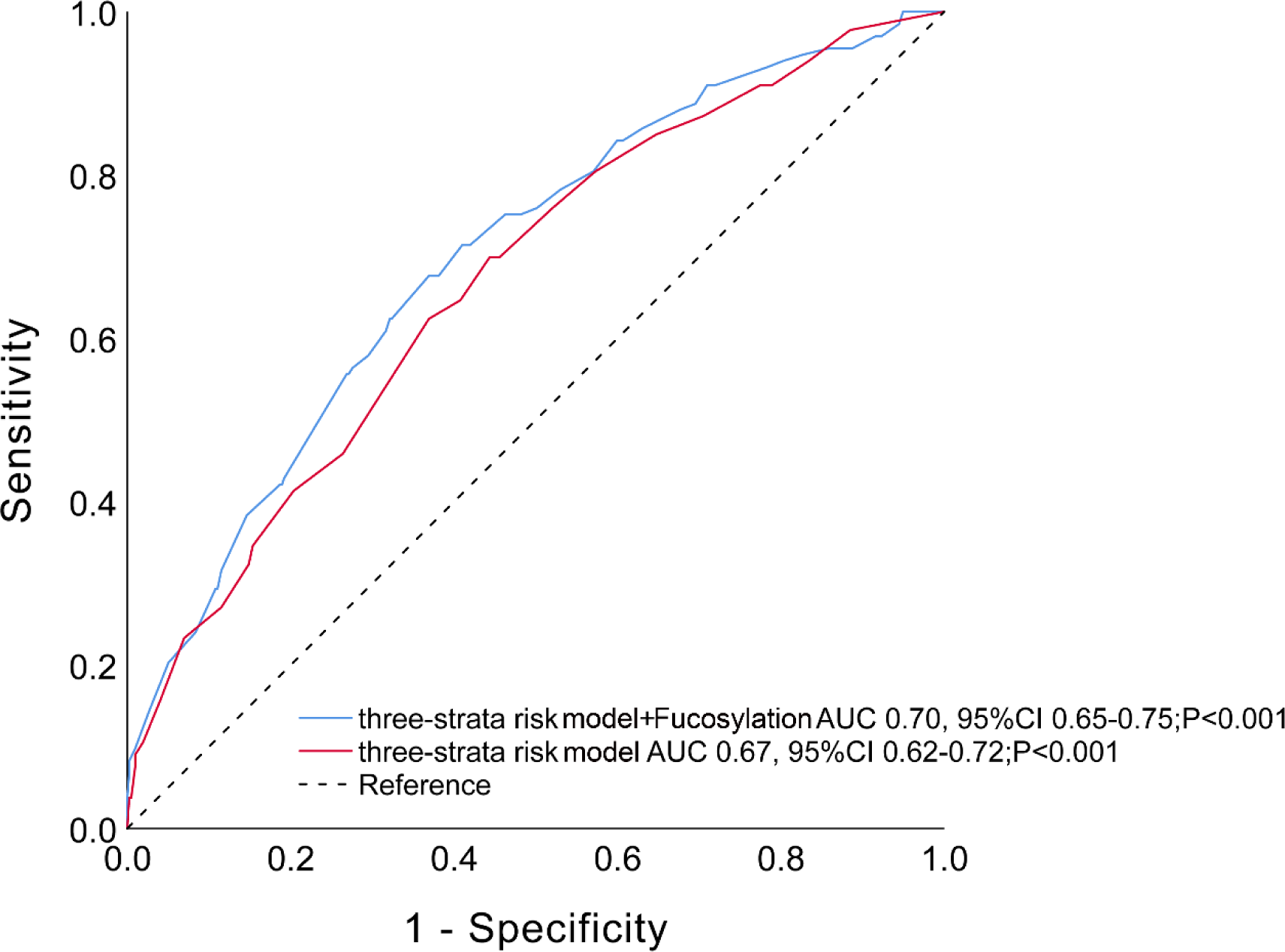
The performance of the established three-strata model (from the SPAHR and COMPERA analyses) before and after the addition of IgG Fucosylation evaluated by receiver-operating characteristic analysis. AUC = area under the curve.

**Figure 6.**
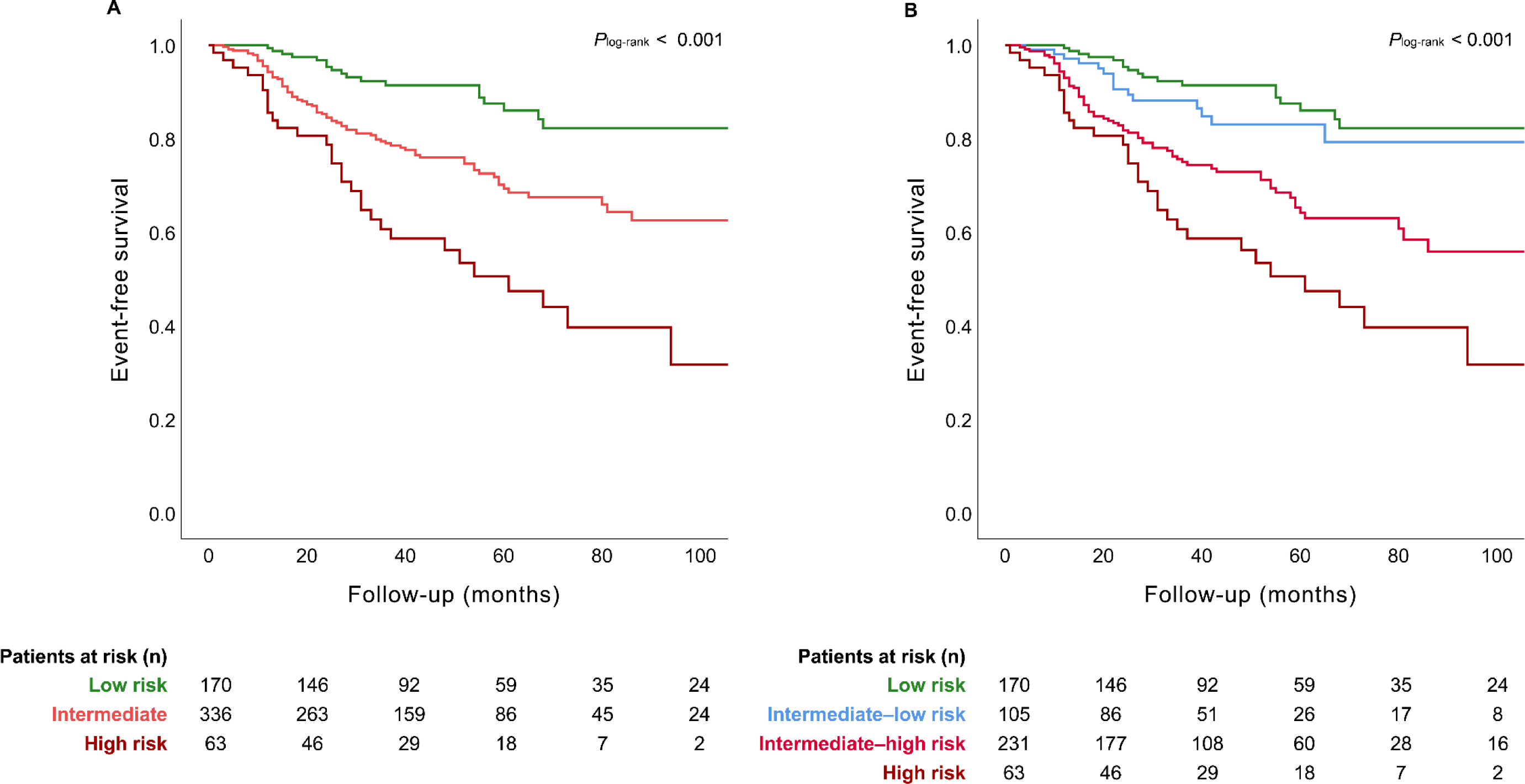
Kaplan-Meier survival curve for all-cause death stratified by the established three-strata model (from the SPAHR and COMPERA analyses) (A) before and (B) after the addition of IgG Fucosylation in patients with pulmonary artery hypertension.

## Discussion

Risk stratification has become an essential part of the management of patients with PAH. The joint guidelines on pulmonary hypertension released in 2015 by the European Society of Cardiology (ESC) and the European Respiratory Society (ERS) introduced a multidimensional risk stratification model integrating 14 variables obtained from nine distinct assessments. According to this model, the risk is categorized into three strata: low, intermediate, or high.^1,2^ However, based on the three-strata model, the majority of patients with PAH were classified as intermediate risk.^30,31^ Despite the development of refined or alternative methods, such as the prognostic risk equation from the French,^33^ SPAHR,^34^ REVEAL,^35^ and COMPERA^36^ Registries, challenges persist regarding the accuracy of stratification and the consistent availability of various clinical parameters during every patient visit. These limitations underscore the need for improvement in the approach to assessing the risk of PAH. In this context, several researchers have attempted to develop new protein biomarkers/panels from blood samples to improve risk assessment.^37–39^ Although these studies have made significant contributions to exploring novel biomarkers for PAH, the latest 2022 ESC/ERS guidelines for PAH continued to recommend NT-proBNP as the exclusive blood biomarker.^40^ Significantly, the studies of glycosylation of plasma proteins as potential biomarkers also shed new light on this field. The roles of plasma IgG N-glycosylation in cardiovascular diseases have been long investigated and discussed. The IgG glycosylation profile was found to be associated with cardiovascular disease risk score and subclinical atherosclerosis.^41^ Additionally, alteration in plasma IgG N-glycosylation profiles was reported to be associated with blood pressure status, suggesting that IgG N-glycans may be potential postgenomic biomarkers for hypertension.^42^ Notably, our recent discovery showcased the utility of IgG Gal-ratio, representing the distribution of IgG galactosylation, in evaluating the inflammatory state in patients with chronic thromboembolic pulmonary hypertension.^24^ These investigations underscored the immense promise of IgG N-glycans as biomarkers for the identification and prognostication of cardiovascular diseases. However, whether IgG N-glycans can be used as prognostic biomarkers for PAH remains to be explored.

To our knowledge, this is the first IgG N-glycomic study in PAH. Applying a robust method based on high-throughput mass spectrometry, we for the first time identified and validated that plasma IgG Fucosylation informs patients’ survival independent of established risk factors. We first screened out the promising candidate prognostic marker IgG Fucosylation in the discovery cohort and validated it in another independent cohort. We then demonstrated that low plasma IgG Fucosylation was robustly associated with an increased risk of death in patients with PAH not only through overall analysis adjusted for potential confounders but also through subgroup analyses. At last, we found that the prognostic IgG Fucosylation improved the clinical risk prediction of the established prognostic risk equation (three-strata model from the SPAHR and COMPERA analyses).^30,31^ This study adopted a rigorous validation scheme as used in other biomarker discovery studies, encompassing the stages of discovery (Beijing cohort), validation (Shanghai cohort), and random sampling analysis (combined cohorts). This comprehensive approach provided robust evidence supporting IgG Fucosylation as a significant prognostic factor in PAH, instilling confidence in the reliability of our findings.

Registries worldwide have consistently indicated a higher prevalence but better prognosis among female patients with PAH.^43^ Interestingly, in the present study, we also observed sex-related differences and low IgG Fucosylation appeared to pose a higher risk in men compared to that in women. Furthermore, in our subgroup analysis based on therapeutic strategies, our results demonstrated that baseline IgG Fucosylation effectively distinguished low-risk and high-risk patients, not only within the treatment-naïve and monotherapy subgroup but also within the dual and triple therapy subgroup. Despite the substantial advancements in the modern era of PAH treatment, a specific subset of patients continues to face a bleak prognosis.

Remarkably, baseline IgG Fucosylation could still discern this subset of patients with adverse outcomes in the context of dual and triple therapy. However, the previously established three-strata model was less effective in predicting outcomes in modern treatment settings compared to monotherapy or treatment-naive subgroups (data not shown). One potential explanation is that the current treatments do not target inflammatory pathways associated with IgG Fucosylation. Although modern dual and triple therapies significantly improve patient survival compared to monotherapy or treatment naïve, this improvement is unrelated to systemic-originated IgG Fucosylation. Baseline IgG Fucosylation can still sensitively predict survival, even more so in the dual and triple therapy groups. These findings suggested IgG Fucosylation may not only be a promising prognostic biomarker but also emerge as a potential novel therapeutic target, distinct from existing targets, for patients with PAH.

In the SPAHR and COMPERA analyses, the majority of patients with PAH were identified as intermediate risk (∼60-70%)^30,31^ and similar results were also obtained in our cohorts using the three-strata model they proposed. Identifying high-risk patients is particularly important in the risk assessment at diagnosis.^44,45^ Discriminating the intermediate-high risk group from the intermediate risk group that makes up the majority of the patients may prompt physicians to initiate more aggressive initial treatment strategies which have far-reaching effects on patients’ long-term survival.^44^ Therefore, a more granular risk stratification is needed for the patients in the intermediate risk group, which is vitally important for therapeutic decisions. Several reports have shown that the use of additional variables improved risk prediction and some groups also proposed the four-strata risk model.^36,46–48^ In the present study, the potential plasma biomarker IgG Fucosylation also achieved promising results with further discrimination within the intermediate risk group. Specifically, the IgG Fucosylation improved discrimination AUC from 0.67 (uninformative test) to 0.70 (moderately accurate test) and reclassification of patients in the intermediate risk group into intermediate-high and intermediate-low risk of the three-strata model, suggesting IgG Fucosylation provided added values in the risk stratification of PAH in combination with the established clinical targets.

Several studies have shown that core fucose on IgG plays a role in the progression of malignant diseases.^49,50^ Functional studies revealed that the lack of core fucose in IgG N-glycome greatly increased the proinflammatory capacity of IgG through increased binding of IgG to FcγRIII and enhanced ADCC.^51,52^ Decreased levels of core Fucosylation of IgG N-glycome coincide with increased inflammation in individuals.^20^ Moreover, in our previous study, we found human plasma IgG N- glycome profiles exhibited a proinflammatory phenotype in chronic thromboembolic pulmonary hypertension and IgG proinflammatory N-glycans may be useful for assessing the occurrence and prognosis of patients with chronic thromboembolic pulmonary hypertension.^24^ In the present study, decreased levels of IgG Fucosylation were found associated with increased mortality of patients with PAH. Therefore, we speculate that inflammation could be a mediator of the prognostic effect of IgG Fucosylation in patients with PAH, but the potential role of changes in IgG Fucosylation in the development and progression of PAH deserves in-depth exploration in future studies.

Circulating BNP or NT-proBNP secreted by cardiac myocytes in response to increased ventricle wall strain^53^ is the only blood biomarker in the established risk calculators. Besides cardiac-originated BNP or NT-proBNP, systemic-originated IgG Fucosylation which is associated with systemic immune and inflammation was found to provide independent values to inform risk stratification in this study. Therefore, plasma biomarkers from different pathways in the pathophysiology of PAH collectively provide more comprehensive information compared to parameters indicating limited mechanisms. On the other hand, though previous studies have reported some new plasma proteins/protein panels may be useful in the risk stratification of PAH, IgG Fucosylation identified in the present study is the first glycomic candidate biomarker for PAH prognosis. Glycomic biomarkers have many advantages over other omic markers. Especially, clinical IgG N-glycosylation analysis does not rely as heavily on mild serum/plasma storage conditions and timely analysis (glycans are even stable at 50 °C for two weeks) as many other omics analyses.^54^ It is worth mentioning that the high-throughput workflow for IgG N-glycome profiling applied in the present study has been widely used in various studies,^24–27,55^ enabling the candidate glycan biomarker to be more convenient for further clinical application.

## Strengths and limitations

A major advantage of this study is that we, for the first time, reliably identified and validated a novel independent prognostic biomarker plasma IgG Fucosylation for PAH using plasma IgG N-glycomics based on relatively large and independent multicenter PAH cohorts with long-term follow-up. Another strength of our study was that the IgG glycan biomarker is systemic-originated and in different pathways of the pathophysiology of PAH from other established risk factors, which provided added values for the prognostication of patients with PAH. Nonetheless, our study has several limitations. First, only idiopathic and heritable PAH were included in our cohorts. The survival in patients with other types of PAH (caused by other associated conditions such as congenital heart disease, connective tissue diseases, and portopulmonary hypertension) may differ significantly from that of patients with idiopathic or heritable PAH. Therefore, applying the conclusions drawn from this study to other categories of PAH requires extreme caution and the prognostic significance of IgG Fucosylation in other subtypes of PAH will be evaluated in future studies. Second, in the present study, we discovered that decreased levels of IgG Fucosylation were independently associated with an increased risk of death in patients with PAH. Though some functional studies have shown that the absence of core fucose in IgG N-glycome affects inflammatory signaling and immune response outcomes, the mechanisms underlying our findings and the potential role of altered IgG Fucosylation in the development and progression of PAH warrant in-depth investigation. Third, we evaluated the prognostic value of baseline plasma IgG N-glycome profiles in PAH. Future investigations should explore the clinical implications of IgG N-glycome variations in patients undergoing diverse stages of treatment. Last, though the sample size of our cohorts was relatively large, the numbers became small in the subgroups of sensitivity analyses and stratified analyses. The accuracy of IgG Fucosylation in prognostication would be increased with an increased number of samples.

## Conclusions

Through a comprehensive scheme, we identified and validated a plasma glycomic prognostic biomarker, which is in different pathophysiological pathways from other established risk factors of PAH, to risk stratify patients with PAH.

## Acknowledgements

### Author Contributions

Conception and design: ZCJ, ZJZ, CL, WHW, JLM, and CYC; Analysis and interpretation of data: ZJZ, CL, JLM, YPZ, DL, TYL, JHL, SJZ, JSM, RNL, and LW; Drafting of the manuscript: ZJZ and CL; Critical revision of the manuscript for important intellectual content: ZCJ, JLM, DL, and YPZ; Final approval of the manuscript: all authors; Statistical expertise: KS; Obtaining research funding: ZCJ and ZJZ; Acquisition of data: LC, ZJZ, JHL, WHW, CYC, and DL.

### Sources of funding

This work was supported by grants from the National Key Research and Development Program of China (2022YFC2703902 and 2022YFC2703901), the CAMS Innovation Fund for Medical Sciences (2021-I2M-1-018), National Natural Science Foundation of China (82241020 and 32371506), Natural Science Foundation of Shanghai (22ZR1452400), Pujiang Talent Program (22PJD064), and National High-Level Hospital Clinical Research Funding (2022-PUMCH-B-099 and 2022-PUMCH-A- 200).

### Disclosures

None

## Supplemental materials

Supplemental methods

Supplemental Figure S1

## Non-standard Abbreviations and Acronyms

6MWD: 6-minute walking distance
AUC: area under the curve
NT-proBNP: N-terminal pro-B-type natriuretic peptide
PAH: pulmonary arterial hypertension
PVR: pulmonary vascular resistance
RAP: right arterial pressure
ROC: receiver-operating characteristic
SvO2: mixed venous oxygen saturation
WHO FC: World Health Organization functional class

## Data Availability

All data produced in the present study are available upon reasonable request to the authors.

