## Supplemental Methods and Figure S1 for "Prognostic value of plasma IgG N-glycome traits in patients with pulmonary arterial hypertension: an observational cohort study"

**Running title**: *Plasma prognostic glycan biomarker for PAH*

Ze-Jian Zhang, PhD[1#]; Chao Liu, MD[2#]; Jie-Ling Ma, MD[2#]; Jing-Si Ma, BS[3]; Jia Wang, BS[4]; Ruo-Nan Li, BS[3]; Dan Lu, MD[5]; Yu-Ping Zhou, MD[2]; Tian-Yu Lian, MD[1]; Si-Jin Zhang, MD[6]; Jing-Hui Li, MD[7]; Lan Wang, MD[8]; Kai Sun, MD[1]; Chun-Yan Cheng, MD[6*]; Wen-Hui Wu[8*]; and Zhi-Cheng Jing, MD[6*]

**Affiliations**

[1]State Key Laboratory of Complex Severe and Rare Diseases, Peking Union Medical College Hospital, Chinese Academy of Medical Sciences and Peking Union Medical College, Beijing 100730, China;

[2]Department of Cardiology, State Key Laboratory of Complex Severe and Rare Diseases, Peking Union Medical College Hospital, Chinese Academy of Medical Sciences and Peking Union Medical College, Beijing 100730, China;

[3]School of Pharmacy, Henan University, Kaifeng 475004, China;

[4]Department of Medical Laboratory, Weifang Medical University, Weifang 261053 China;

[5]Cardiac Department, Aerospace Center Hospital, Peking University Aerospace School of Clinical Medicine, Beijing 100049, China;

[6]Department of Cardiology, Guangdong Institute of Cardiovascular Diseases,

Guangdong Provincial People's Hospital, Guangdong Academy of Medical Sciences, Southern Medical University, Guangzhou 510080, China;

[7]State Key Laboratory of Cardiovascular Disease and FuWai Hospital, Chinese Academy of Medical Sciences and Peking Union Medical College, Beijing 100037, China;

[8]Department of Cardio-Pulmonary Circulation, Shanghai Pulmonary Hospital, School of Medicine, Tongji University, Shanghai 200433, China.

[#]These authors contributed equally to this work as co-first authors.

[*]These authors contributed equally to this study as co-corresponding authors.

**Address for Correspondence:**

Zhi-Cheng Jing, MD, PhD

Department of Cardiology, Guangdong Institute of Cardiovascular Diseases, Guangdong Provincial People's Hospital, Guangdong Academy of Medical Sciences, Southern Medical University, No.106, Zhongshan 2nd Road, Guangzhou 510080, China.

Wen-Hui Wu, MD

Department of Cardio-Pulmonary Circulation, Shanghai Pulmonary Hospital, School of Medicine, Tongji University, Shanghai 200433, China, No.507, Zhengmin Road, Shanghai 200433, China.

Chun-Yan Cheng, MD

Department of Cardiology, Guangdong Institute of Cardiovascular Diseases, Guangdong Provincial People's Hospital, Guangdong Academy of Medical Sciences, Southern Medical University, No.106, Zhongshan 2nd Road, Guangzhou 510080, China.

**Supplemental Methods**

**Detailed procedures for mass spectrometric IgG N-glycome profiling**

The plasma samples of the two cohorts were prepared for IgG N-glycosylation profiling as previously described[1-3]. For this, IgG was isolated from plasma samples using a 96-well Protein A Spin Plate (Thermo Fisher Scientific) according to a previously published method[1-3]. Briefly, the Protein A Spin Plate was first equilibrated by adding 300 μL of Binding Buffer (Thermo Fisher Scientific, 0.5L) to each well. Then, 140 μL of the diluted plasma (1:1, by Binding Buffer) from each participant was added to the preconditioned Spin Plate, followed by incubation for 1 h on a plate shaker to achieve IgG binding. Subsequently, the Spin plate was washed five times with 300 μL of Binding Buffer to wash away non-IgG proteins. Last, IgG was eluted from the Spin Plate by Elution Buffer (Thermo Fisher Scientific, 0.5L). The purity of the eluted IgG was confirmed by sodium dodecyl sulfate-polyacrylamide gel electrophoresis. The IgG solution was stored at −20 °C until use. IgG N-glycan release was performed according to our previous study1-3. Briefly, 50 μL of denatured IgG solution was mixed with the release mixture containing 1 U PNGase F (New England Biolabs), and then incubated for 12 h at 37 °C. IgG N-glycan purification was performed using a porous graphitized carbon (PGC)-containing 96-well plate[1,3]. The PGC plate was prewetted twice with 200 μL (for each well) of 0.1% trifluoroacetic acid (TFA, Sigma-Aldrich) (v/v) in 80% acetonitrile (ACN) (v/v) and conditioned twice with 200 μL of 0.1% TFA (v/v) in $H_2O$. Next, the enzymatic digest was applied to the PGC plate three times. The PGC plate was washed twice with $H_2O$. The enriched and purified glycans were finally eluted with

0.05% TFA(v/v) in 25% ACN (v/v). Matrix-assisted laser desorption/ionization time of flight mass spectrometry (MALDI-TOF-MS) measurements were performed as previously described[3,4]. In short, 1 μL of glycans solution was spotted with 1 μL of the matrix (containing 5 mg/mL sDHB) onto a target plate (MTP AnchorChip 384BC, Bruker Daltonics) and the spots were left to dry for 2 h. The samples were then recrystallized with ethanol. A Bruker rapifleX MALDI-TOF-MS with a Smartbeam 3D laser recorded the spectra in reflection positive mode in the m/z range from 1000 to 3500. The system was operated by flexControl (version 4.0) software (Bruker Daltonics). 5K laser shots were accumulated per spot in a complete sample random walk pattern with 100 shots per raster spot at a laser frequency of 5000 Hz. The processing of MALDI-TOF-MS raw data and IgG N-glycan identification was according to our previous work[2-4]. Briefly, raw MS data were baseline-subtracted, smoothed, and transformed to XY files using flexAnalysis (version 3.4) software (Bruker Daltonics). The XY profiles were re-calibrated and the intensities for the N-glycan structure signals were extracted as background-corrected relative areas for each spectrum using MassyTools (version 0.1.8.1). Detailed information on glycan identification was described in our published work[2-4]. Of note, if multiple cation adduct signals ($[M + Na]^+$ and $[M + K]^+$) of one glycan structure were present simultaneously, all the adducts should be considered for the quantification.

### Supplemental Figure

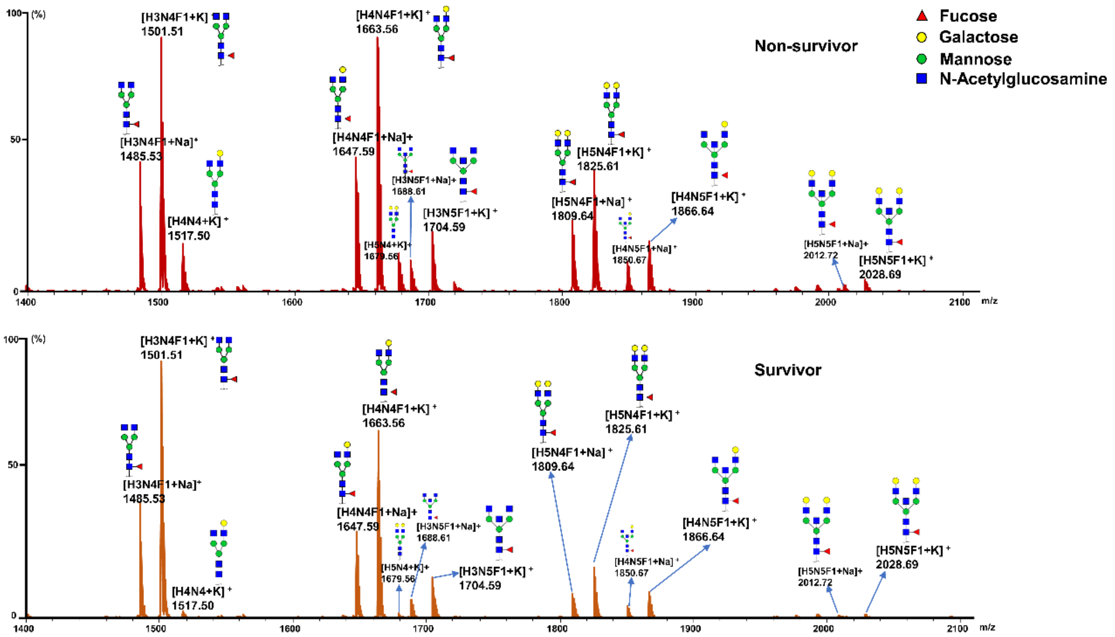


**Figure S1.** Typical annotated matrix-assisted laser desorption/ionization time of flight mass spectrometry spectra of plasma IgG N-glycome acquired from the survivor and non-survivor patients with pulmonary arterial hypertension.
